# AI-detected tumor-infiltrating lymphocytes for predicting outcomes in anti-PD1 based treated melanoma

**DOI:** 10.1101/2025.05.28.25328410

**Authors:** Mark Schuiveling, Isabella A.J. van Duin, Laurens ter Maat, Janneke C. van der Weerd, Rik J. Verheijden, Franchette van den Berkmortel, Christian U. Blank, Gerben E. Breimer, Femke H. Burgers, Marye Boers-Sonderen, Alfons J. M. van den Eertwegh, Jan Willem B. de Groot, John B.A.G. Haanen, Geke A.P. Hospers, Ellen Kapiteijn, Djura Piersma, Gerard Vreugdenhil, Hans Westgeest, Anne M.R. Schrader, Josien Pluim, Paul J. van Diest, Mitko Veta, Karijn P.M. Suijkerbuijk, Willeke Blokx

## Abstract

**Importance:** Easy and accessible biomarkers to predict response to immune checkpoint inhibition (ICI)-treated melanoma are limited.

**Objective:** To evaluate artificial intelligence (AI) detected tumor-infiltrating lymphocytes (TILs) on pretreatment melanoma metastases as a biomarker for response and survival in ICI-treated patients.

**Design:** Multicenter cohort study including patients with advanced melanoma treated with first-line anti-PD1 ± anti-CTLA4 between 2016 and 2023. Median follow-up was 36.3 months.

**Setting:** 11 melanoma treatment centers in the Netherlands.

**Participants:** 1,202 patients with advanced cutaneous melanoma.

**Exposure:** All patients received first-line anti-PD1 ± anti-CTLA4.

**Main Outcome(s) and Measure(s):** The percentage of TILs inside manually annotated tumor area in H&E stained pretreatment metastases was determined using the Hover-NeXt model trained and evaluated on an independent melanoma dataset containing 166,718 pathologist-verified manually annotated cells. The primary outcome was objective response rate (ORR); secondary outcomes were progression-free survival (PFS) and overall survival (OS). Correlation with manual TILs, scored according to the guidelines stated by the immune-oncology working group, was evaluated with Spearman correlation coefficients. Logistic regression and Cox proportional regression were conducted, adjusted for age, sex, disease stage, ICI type, BRAF status, brain metastases, LDH level, and performance status.

**Results:** Metastatic melanoma specimens were available for 1,202 patients, of whom 423 received combination therapy. Median TIL percentage was 9.9% (range 0.3% - 69.4%). A 10% increase in TILs was associated with increased ORR (adjusted OR 1.40 [95% 1.23-1.59]), PFS (adjusted HR 0.85 [95% CI 0.79 – 0.92]) and OS (adjusted HR 0.83 [95% CI 0.76 – 0.91]. Results were consistent for both patients treated with anti-PD1 monotherapy and combination treatment with anti-PD1 plus anti-CTLA4. When comparing manual TIL scoring with AI-detected TILs, associations with response and survival were consistently stronger for AI-detected TILs.

**Conclusions and Relevance:** In patients with advanced melanoma, higher levels of AI-detected TILs on pre-treatment H&E slides were independently associated with improved ICI response and survival. Given the accessibility of TIL scoring on routine histology, TILs may serve as a predictive biomarker for ICI outcomes. To facilitate broader validation, the Hover-NeXt architecture and model weights are publicly available.

**Key points:** **Question:** What is the predictive value of artificial intelligence-detected tumor-infiltrating lymphocytes (TILs) for clinical outcomes in patients with advanced melanoma receiving first-line immune checkpoint inhibition?

**Findings:** In this multicenter cohort of 1202 patients, TILs in pretreatment metastases were quantified using a melanoma-specific publicly available AI model trained on an independent dataset. A 10% increase was associated with response (aOR 1.40 [95% CI 1.23–1.59]), progression-free survival (aHR 0.85 [95% CI 0.79–0.92]), and overall survival (aHR 0.83 [95% CI 0.76–0.91]). Associations were independent of clinical predictors.

**Meaning:** AI-detected TILs in pretreatment melanoma metastases independently correlate with response and survival.

## Introduction

Immune checkpoint inhibition (ICI) has significantly improved the prognosis for patients with advanced cutaneous melanoma. In the Checkmate 067 trial, ten-year overall survival (OS) rates of 37% for anti-PD1 monotherapy and 43% for combination therapy with anti-CTLA4 have been reported.^1^ However, significant challenges remain. Despite not all patients responding to therapy, many still experience side effects, with severe toxicity occurring in up to 60% of patients receiving combination treatment ^2,3^. Furthermore, ICIs place a substantial financial burden on the healthcare system, with average medication costs per patient reaching €60,000 for monotherapy and €117,000 for combination therapy in The Netherlands.^4,5^ Therefore, there is a need for predictive biomarkers that can identify patients most likely to benefit from ICI treatment.

Our previous research identified tumor-infiltrating lymphocytes (TILs) as such a potential biomarker. In a cohort of 650 patients, we showed that the percentage of intra-tumoral stroma occupied by TILs, which is the scoring method defined by the International TILs Working Group^6^, was associated with both treatment response and survival outcomes.^7^ However, it also became clear that manual scoring is hindered by substantial interobserver variability.^7–9^

Artificial intelligence (AI) offers a scalable solution by enabling reproducible and quantitative assessment of TILs on routine H&E slides. While AI-based TIL quantification has demonstrated prognostic value in multiple tumor types ^10–12^, evidence in melanoma remains inconsistent. Prior studies in advanced melanoma cohorts reported conflicting associations between AI-detected TILs and clinical outcomes, likely due to methodological variability, small cohort sizes, and limited model generalizability ^13–16^.

To address these challenges, we developed an open-source, melanoma-specific AI model to detect TILs on H&E-stained slides ^16^. In this study, we applied this model to pre-treatment metastatic melanoma samples to evaluate whether AI-detected TILs are associated with ICI response and survival in a real-world cohort of patients with advanced disease.

## Methods

### TIL model and developmental cohort

The TIL detection model used in this study was trained and evaluated on an independent developmental cohort consisting of 155 unique primary and 197 unique metastatic melanoma samples. This dataset included 161,835 nuclei annotations verified by a dermatopathologist (author W.A.M.B.). Part of the developmental dataset, along with the cell detection model and the melanoma-specific model weights, are publicly available.^17,18^ Further details on the construction of the developmental cohort and the model training process are provided in the data supplements.^16,19,20^

### Clinical cohort

Patients were retrospectively identified from eleven melanoma treatment centers using high quality prospectively collected registry data.^21^ Patients were included if above 18 years of age and treated with first-line anti-PD1 ± anti-CTLA4 for unresectable stage IIIC or stage IV melanoma after January 1, 2016 until the 1^st^ of January 2023. The patient’s stage of disease was based on the 8^th^ edition of the AJCC melanoma staging system.^22^

For application of the model to the patient cohort, the tumor area excluding large areas of stroma and necrosis was delineated on the digitized specimen. This was performed manually (by authors J.v.d.W. and M.S., blinded for the outcome) under the supervision of a dermatopathologist (author W.A.M.B.).^23–25^ The TIL score was defined as the percentage of AI-detected TILs among all cells detected within the annotated tumor area. A visual representation of the TIL scoring is shown in Figure 1 and an example output of the model is visualized in Supplementary Figure 1.

**Figure 1.**
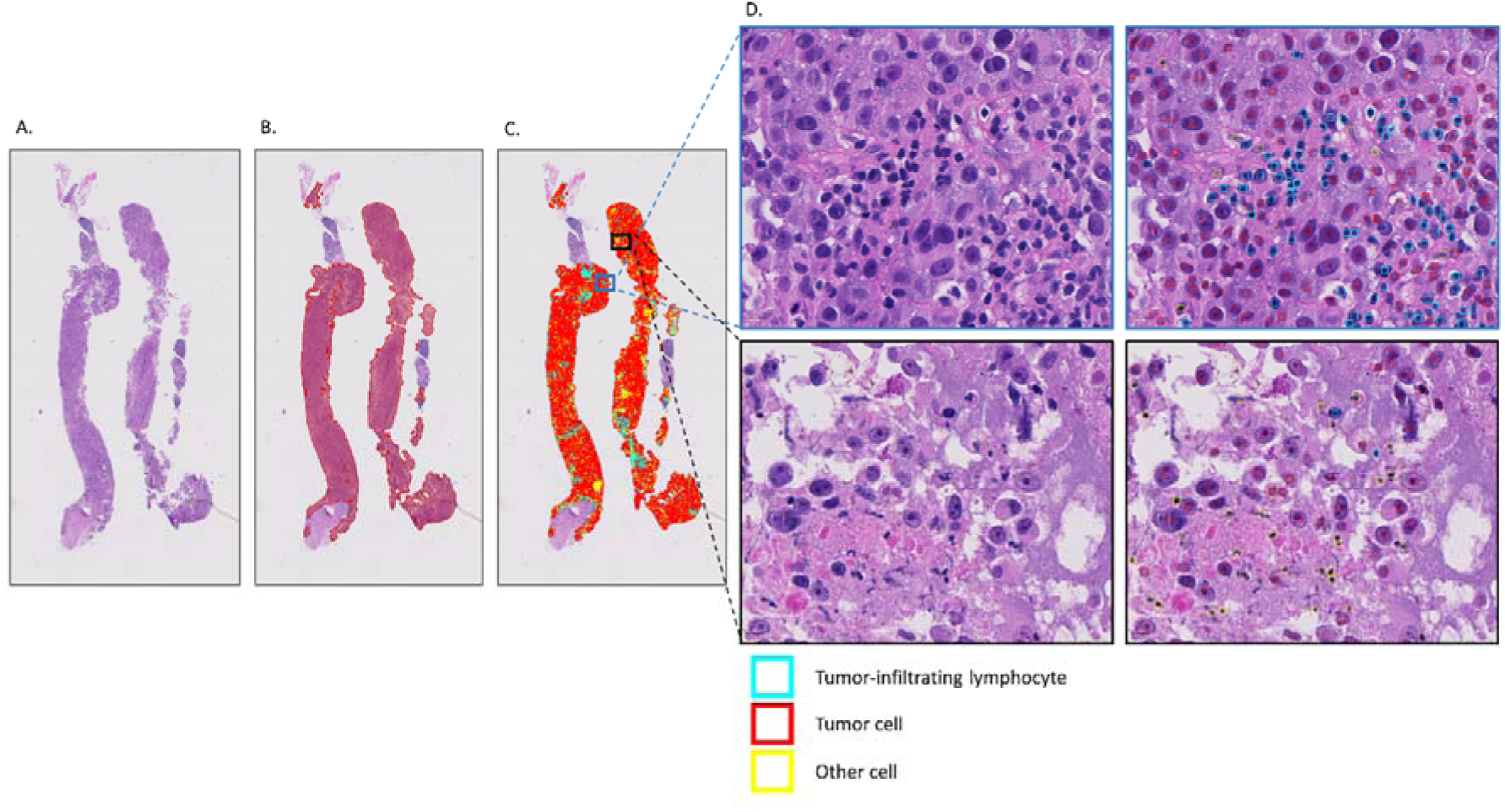
Shown in the panels are a digitized whole slide image of a melanoma biopsy taken from a metastasis in a lymph node (A), the manually annotated tumor area (B), the output of the cell detection model filtered to the annotated tumor area (C), and a zoomed-in view of the cell detection results (D) with one zoomed in region with annotated tumor-infiltrating lymphocytes and one zoomed in region with tumor cells and apoptotic cells segmented as other. From all cells present, the percentage of tumor-infiltrating lymphocytes for the tumor area is calculated as:

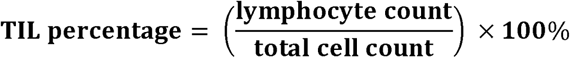

### Slide selection

Slides were collected from 30 different hospital archives through PALGA, the Dutch nationwide registry of histo- and cytopathology.^26^ For each patient, a single representative H&E-stained slide was selected. In cases with multiple specimens, the largest available specimen before therapy initiation was used. Curation and selection were done by four authors (M.S., I.A.J.v.D., L.t.M., and J.v.d.W.) under supervision of two experienced pathologists (W.A.M.B. and P.J.v.D.). All selected slides were scanned with a Nanozoomer XR C12000-21/-22 (Hamamatsu Photonics, Hamamatsu, Shizuoka, Japan) at 40× magnification with a resolution of 0.23 µm per pixel. The anatomic location of the metastatic lesion was derived from the pathology report if described or assessed visually on the slide when possible (in case of presence of pre-existing normal tissue of the organ). Determination of the nature of the specimen type (resection or biopsy) was visually assessed.

### Manual TIL classification

Manual TIL scoring was performed by authors I.A.J.v.D. and M.S., under the supervision of two experienced pathologists (W.A.M.B. and P.J.v.D.). TILs were scored using the semi-quantitative method proposed by the International Immuno-Oncology Biomarker Working Group. This method estimates the percentage of TILs present within the intra-tumoral stromal area.^6^

### Clinical variables

Clinical data included age at treatment initiation, sex, ICI type, World Health Organization (WHO) performance status, BRAF V600E mutation status, serum lactate dehydrogenase (LDH) level, AJCC disease stage, and presence of symptomatic brain metastases. We incorporated the variable ‘presence of symptomatic brain metastases’ in the variable ‘stage of disease’: M1D stage was subcategorized either as ‘M1D-non-symptomatic’ or ‘M1D-symptomatic’. LDH levels were categorized as normal, 1-2 times the upper limit of normal (ULN) or twice the upper limit of normal. WHO performance status was categorized as 0, 1, or 2 and above.

### Outcome measures

Response evaluation was determined by the treating physician and was based on the Response Evaluation Criteria in Solid Tumors, version 1.1, with melanoma-related death before first response assessment classified as progressive disease.^27^ Primary outcome was objective response, defined as the best overall response (partial or complete response) within 6 months. Secondary outcomes were progression-free survival (PFS) and overall survival (OS).

### Missing data

To account for missing covariate data, we used multiple imputation by chained equations (MICE) under the assumption that the data were Missing At Random (MAR). The mice package (version 3.15) in R was used to generate 100 imputed datasets over 20 iterations, which is by default more than the proposed minimum of datasets (the percentage of incomplete cases). Convergence of the imputation model was verified visually using trace plots.

### Statistical analysis

Continuous variables were summarized using median and range or interquartile range (IQR), and categorical variables with frequencies and percentages. Associations between continuous variables and response were assessed using the Mann-Whitney U test, assuming non-normal distribution. Correlations were evaluated with Spearman correlation coefficients and visualized using scatter plots. Median follow-up was estimated using the reversed Kaplan-Meier method. PFS and OS were visualized with Kaplan-Meier curves, and differences between groups were assessed using the logrank test.

Univariable and multivariable analyses were conducted using logistic regression and Cox proportional hazards regression. Model assumptions were visually checked and showed no violations. Multivariable analyses were performed on imputed datasets, with results pooled using Rubin’s rules. Unless stated otherwise, all estimates are based on these pooled analyses. Statistical analyses were performed using R (version 4.2.2; survival package version 3.5.0).

## Results

### AI-detected TILs model performance

Our developed cell detection model achieved an F_1_ score of 0.79 [95% CI 0.77 – 0.80] for TIL detection, closely approaching the interobserver agreement on a subset of the developmental dataset (F_1_ score of 0.83 [95% CI 0.79 – 0.88]). Visual inspection revealed no remaining major misclassifications. Examples of the model’s output on the clinical dataset are displayed in Supplementary Figure 1.

### Patient characteristics and outcomes

Of the 1935 eligible patients, 44 were excluded due to missing survival outcome data. From the remaining 1891 patients, metastatic specimens with viable tumor area were available for 1202 patients (Supplementary Figure 2). Patient characteristics are shown in Table 1 and compared well to those of excluded patients (Supplementary Table 1). Most patients were male, had non-elevated LDH levels, and were above 65 years of age. Lymph nodes and skin were identified as the most common site of origin for the metastatic specimens. In patients treated with combination therapy, the analyzed specimen was more often a brain or liver metastasis.

**Table 1.**
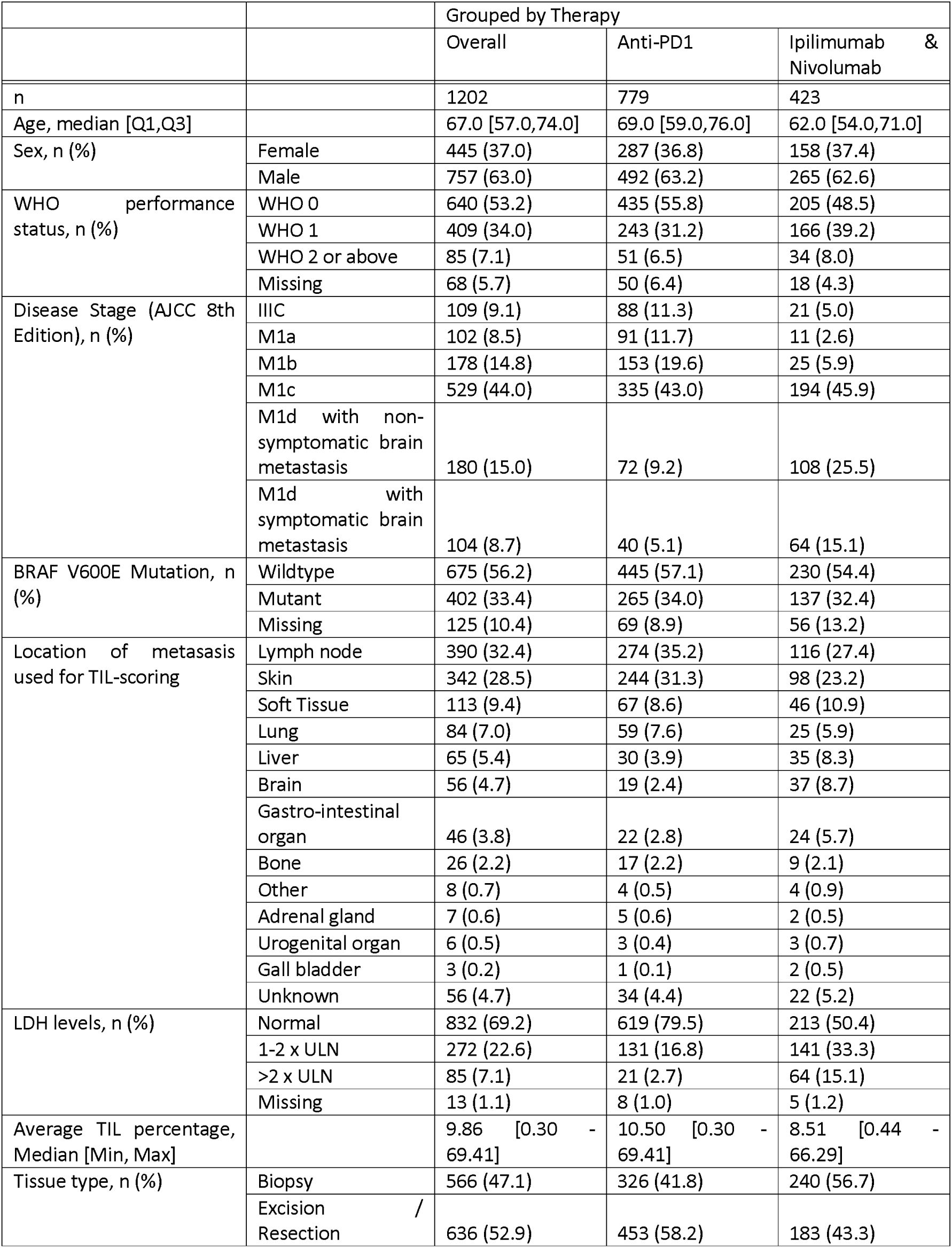
Patient characteristics of 1202 ICI-treated patients with pre-treatment metastatic specimen available. Abbreviations: IQI, interquartile interval; LDH, Lactate dehydrogenase; TIL, tumor-infiltrating lymphocytes; ULN, upper limit of normal; WHO, World Health Organization.

|  |  | Grouped by Therapy |  |  |
| --- | --- | --- | --- | --- |
|  |  | Overall | Anti-PD1 | Ipilimumab & Nivolumab |
| n |  | 1202 | 779 | 423 |
| Age, median [Q1,Q3] |  | 67.0 [57.0,74.0] | 69.0 [59.0,76.0] | 62.0 [54.0,71.0] |
| Sex, n (%) | Female | 445 (37.0) | 287 (36.8) | 158 (37.4) |
|  | Male | 757 (63.0) | 492 (63.2) | 265 (62.6) |
| WHO performance status, n (%) | WHO 0 | 640 (53.2) | 435 (55.8) | 205 (48.5) |
|  | WHO 1 | 409 (34.0) | 243 (31.2) | 166 (39.2) |
|  | WHO 2 or above | 85 (7.1) | 51 (6.5) | 34 (8.0) |
|  | Missing | 68 (5.7) | 50 (6.4) | 18 (4.3) |
| Disease Stage (AJCC 8th Edition), n (%) | IIIC | 109 (9.1) | 88 (11.3) | 21 (5.0) |
|  | M1a | 102 (8.5) | 91 (11.7) | 11 (2.6) |
|  | M1b | 178 (14.8) | 153 (19.6) | 25 (5.9) |
|  | M1c | 529 (44.0) | 335 (43.0) | 194 (45.9) |
|  | M1d with non-symptomatic brain metastasis | 180 (15.0) | 72 (9.2) | 108 (25.5) |
|  | M1d with symptomatic brain metastasis | 104 (8.7) | 40 (5.1) | 64 (15.1) |
| BRAF V600E Mutation, n (%) | Wildtype | 675 (56.2) | 445 (57.1) | 230 (54.4) |
|  | Mutant | 402 (33.4) | 265 (34.0) | 137 (32.4) |
|  | Missing | 125 (10.4) | 69 (8.9) | 56 (13.2) |
| Location of metasasis used for TIL-scoring | Lymph node | 390 (32.4) | 274 (35.2) | 116 (27.4) |
|  | Skin | 342 (28.5) | 244 (31.3) | 98 (23.2) |
|  | Soft Tissue | 113 (9.4) | 67 (8.6) | 46 (10.9) |
|  | Lung | 84 (7.0) | 59 (7.6) | 25 (5.9) |
|  | Liver | 65 (5.4) | 30 (3.9) | 35 (8.3) |
|  | Brain | 56 (4.7) | 19 (2.4) | 37 (8.7) |
|  | Gastro-intestinal organ | 46 (3.8) | 22 (2.8) | 24 (5.7) |
|  | Bone | 26 (2.2) | 17 (2.2) | 9 (2.1) |
|  | Other | 8 (0.7) | 4 (0.5) | 4 (0.9) |
|  | Adrenal gland | 7 (0.6) | 5 (0.6) | 2 (0.5) |
|  | Urogenital organ | 6 (0.5) | 3 (0.4) | 3 (0.7) |
|  | Gall bladder | 3 (0.2) | 1 (0.1) | 2 (0.5) |
|  | Unknown | 56 (4.7) | 34 (4.4) | 22 (5.2) |
| LDH levels, n (%) | Normal | 832 (69.2) | 619 (79.5) | 213 (50.4) |
|  | 1-2 x ULN | 272 (22.6) | 131 (16.8) | 141 (33.3) |
|  | >2 x ULN | 85 (7.1) | 21 (2.7) | 64 (15.1) |
|  | Missing | 13 (1.1) | 8 (1.0) | 5 (1.2) |
| Average TIL percentage, Median [Min, Max] |  | 9.86 [0.30 - 69.41] | 10.50 [0.30 - 69.41] | 8.51 [0.44 - 66.29] |
| Tissue type, n (%) | Biopsy | 566 (47.1) | 326 (41.8) | 240 (56.7) |
|  | Excision / Resection | 636 (52.9) | 453 (58.2) | 183 (43.3) |

The median follow-up was 36.3 months with a median PFS of 9.0 months and a median OS of 35.0 months. The objective response rate (ORR) to ICI was 56.7%.

The median average TIL percentage was 9.9 [range 0.3% - 69.4%]. In patients receiving anti-PD1 therapy, the median average percentage was 10.5 [range 0.3% - 69.4%], while in those treated with combination therapy, it was 8.6% [range 0.4% - 66.3%]. Higher TIL percentages were observed in lymph node (12.8% [range 1.2% - 69.4%]) and lung (11.3% [range 0.4% - 53.5%]) metastases, while liver metastases had lower TIL levels (7.3% [range 0.8% - 26.9%]; Supplementary Table 2).

### AI-detected TILs and response

Patients who responded to therapy had significantly higher median average TIL levels (10.9% [IQR 6.1% - 18.6%] when compared to non-responders 8.3% [IQR 4.6% - 14.8%] *P*<.001). When stratified to treatment type, this effect remained significant in both patients treated with anti-PD1 monotherapy (12.4% [IQR 6.9% - 20.7%] vs 8.6% [IQR 5.0% - 16.3%] *P* <,001), and in patients treated with anti-PD1 and anti-CTLA4 combination therapy (9.5% [IQR 5.4% - 14.9%]%] vs 7.6% [IQR 4.2% - 13.6%]] *P* =.049, Supplementary Figure 3).

Upon dividing TIL levels into tertiles, ORR was 46.4% in the lowest tertile, increasing to 56.4% in the middle and 63.7% in the highest tertile. In the anti-PD1 monotherapy subgroup, ORRs increased from 43.8% to 55.8% and 64.9% across the tertiles, whereas in the combination therapy group, the rates were 50.4%, 61.7%, and 58.2% in the low, middle, and highest tertiles, respectively (Table 2). Cut-offs for the tertiles are 6.9% and 13.1% for all patients, 7.3% and 14.3% for the anti-PD1 treated patients and 6.1% and 11.7% for the patients treated with anti-PD1 combined with anti-CTLA4. Among the 5% of patients with the lowest TIL levels (cut-off1≤12.3%), the ORR was 40.7%, compared to 74.6% in the highest 5% (cut-off1≥135.4%).

**Table 2.** Association of AI-detected TILs with response, progression free survival and overall survival in 1202 patients treated with immune checkpoint inhibition. Results are also shown stratified for 779 patients treated with anti-PD1 monotherapy and 423 patients treated with anti-PD1 + anti-CTLA4. Abbreviations: ICI, immune checkpoint inhibition; CI, confidence interval; OR, odds ratio; HR, hazard ratio; ORR, objective response rate; PFS, progression free survival; OS, overall survival

| Outcome | ICI type | ORR % / median months [95% CI] | ORR % / median months [95% CI] | ORR % / median months [95% CI] | Univariable OR / HR [95% CI] | Multivariable OR / HR [95% CI] |
| --- | --- | --- | --- | --- | --- | --- |
| Group |  | Lowest TIL Tertile | Middle TIL Tertile | Highest TIL Tertile |  |  |
| ORR | All | 46.4% [41.6 - 51.3] | 56.4% [51.5 - 61.1] | 63.7% [58.9 - 68.3] | 1.37 [1.21 – 1.54] | 1.40 [1.23 – 1.59] |
|  | Anti-PD1 | 43.8% [37.9 - 49.9] | 55.8% [49.7 - 61.7] | 64.9% [58.9 - 70.4] | 1.42 [1.23 – 1.65] | 1.44 [1.23 – 1.67] |
|  | Anti-PD1 + Anti-CTLA4 | 50.4% [42.2 - 58.5] | 61.7% [53.5 - 69.3] | 58.2% [49.9 - 66.0] | 1.28 [1.03 – 1.62] | 1.36 [1.07 – 1.73] |
| PFS | All | 5.3 [4.5 - 6.5] | 9.7 [8.1 - 14.0] | 15.6 [10.9 - 20.1] | 0.85 [0.79 – 0.91] | 0.85 [0.79 – 0.92] |
|  | Anti-PD1 | 5.2 [3.7 - 7.0] | 12.8 [8.6 - 15.4] | 15.6 [10.0 - 20.2] | 0.84 [0.77 – 0.91] | 0.85 [0.78 – 0.93] |
|  | Anti-PD1 + Anti-CTLA4 | 5.3 [3.5 - 8.1] | 10.1 [6.4 - 34.7] | 12.0 [6.2 - 28.4] | 0.88 [0.77 – 1.01] | 0.83 [0.73 – 0.96] |
| OS | All | 21.4 [18.5 - 30.8] | 35.9 [28.8 - 58.4] | 49.6 [41.3 - NA] | 0.81 [0.74 – 0.88] | 0.83 [0.76 – 0.91] |
|  | Anti-PD1 | 21.3 [18.5 - 30.5] | 34.9 [28.4 - 58.4] | 49.8 [41.3 - NA] | 0.79 [0.72 – 0.88] | 0.83 [0.75 – 0.92] |
|  | Anti-PD1 + Anti-CTLA4 | 21.1 [13.9 - 41.3] | 77.8 [28.8 - NA] | 49.6 [22.2 - NA] | 0.85 [0.72 – 1.00] | 0.83 [0.70 – 0.97] |

In univariable analysis, a 10% increase in baseline TIL percentage was associated with a greater likelihood of response (OR 1.37 [95% CI 1.21–1.54]). This association remained significant in multivariable analysis after adjusting for established clinical predictors: age, sex, disease stage, WHO performance status, LDH levels, BRAF V600 mutation status, presence of symptomatic brain metastases, and type of therapy (adjusted OR [aOR] 1.40 [95% CI 1.23–1.59]; Table 2 and Supplementary Table 3). When stratified by therapy type, TILs were predictive of response in both anti-PD1 monotherapy (aOR 1.44 [95% CI 1.23–1.67]) and combination therapy with anti-PD1 and anti-CTLA4 (aOR 1.36 [95% CI 1.07–1.73]; Table 2).

### AI-detected TILs and survival

TILs were associated with an improved PFS and OS. Patients in the highest tertile had a median PFS of 15.6 months, while those in the middle and lowest tertiles had PFS of 9.7 and 5.3 months, respectively (*P*<.001). OS was 49.6 months in the highest tertile, compared to 35.9 and 21.4 months in the middle and lowest tertiles (*P*<.001). Results were similar for patients treated with anti-PD1 monotherapy, however in patients treated with anti-PD1 + anti-CTLA4 combination therapy there was no clear separation between the middle and highest tertile in both the OS and PFS analysis (*P*=0.02 for OS and *P*=0.01 for PFS, Figure 2).

**Figure 2.**
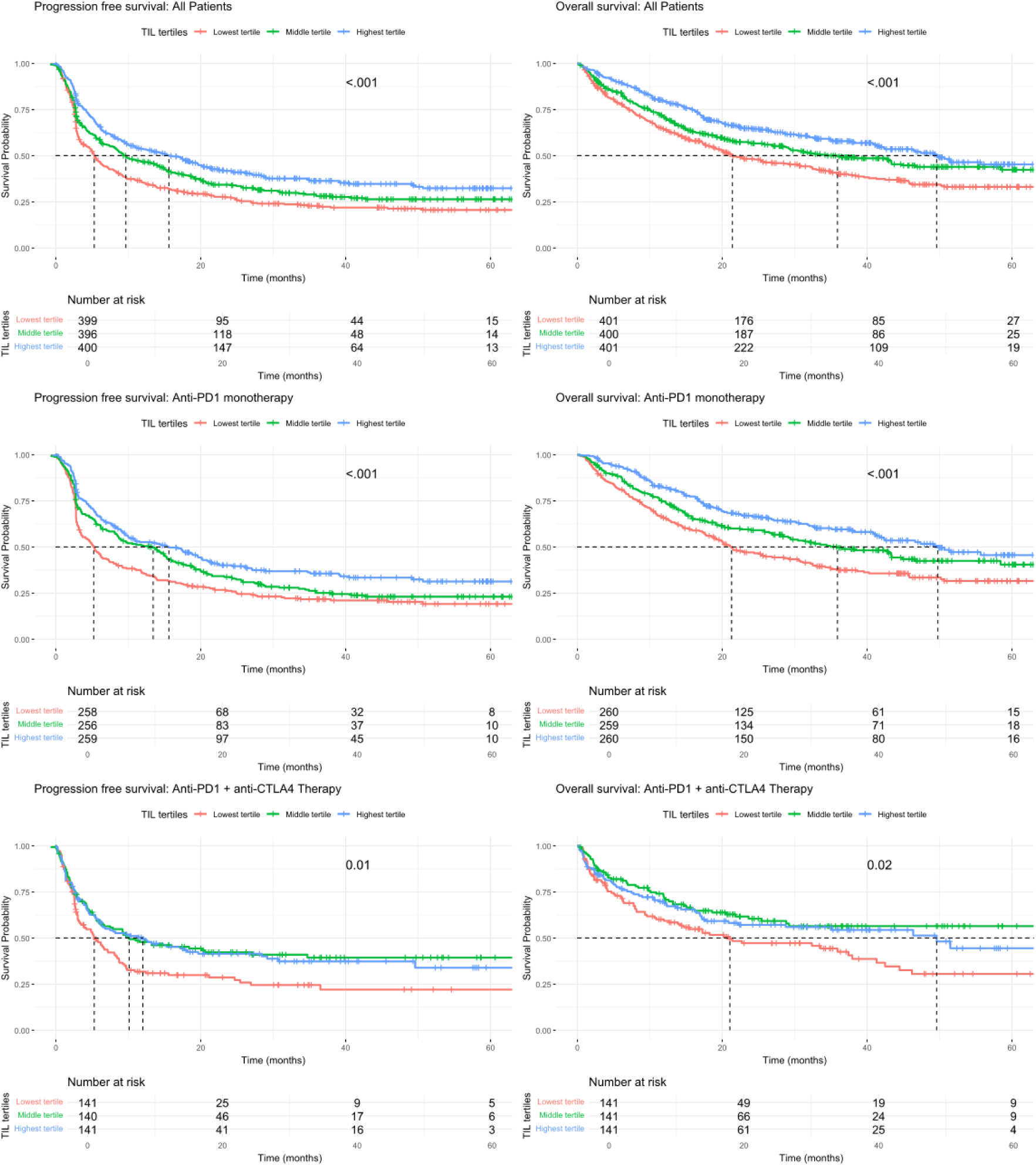
Kaplan-Meier curves for progression-free survival (PFS) and overall survival (OS) grouped by low, intermediate, and high levels of AI-detected tumor-infiltrating lymphocytes in pre-treatment metastatic specimens of patients with advanced melanoma. Curves are shown for all patients (n = 1202, panels A and B), those treated with anti-PD1 monotherapy (n = 779, panels C and D), and those receiving combination therapy (n = 423, panels E and F). PFS data were missing for 7 patients. Differences between groups were assessed using log-rank tests.

When using TILs as a continuous variable, a 10% increase in baseline TIL percentage was associated with a decreased risk of progression (HR 0.85, 95% CI [0.79–0.91]) and death (HR 0.81, 95% CI [0.74– 0.88]). This effect remained significant in multivariable analysis (adjusted HR 0.85 [95% CI 0.79–0.92] for PFS and 0.83 [95% CI 0.76–0.91] for OS; Table 2 and Supplementary Table 3). When stratified based on treatment, similar results were seen for both patients treated with anti-PD1 monotherapy and anti-PD1 combined with anti-CTLA4 (Table 2).

### Biopsy versus surgical excision

TILs remained predictive in both surgical resections and biopsies, with an odds ratio (OR) of 1.32 [95% CI 1.14–1.52] for resections and 1.54 [95% CI 1.24–1.91] for biopsies. Median average AI-detected TIL percentage was 9.4% [IQR 5.3% - 15.2%] in biopsies, compared to 10.4% [IQR 5.6% - 18.6%] in resections. The median annotated tumor area was larger in resections (96.3 mm² [IQR 35.3 – 197.4]) than in biopsies (11.3 mm² [IQR 5.4 – 24.0]).

### Correlation with manual TIL scoring

AI-detected TILs showed a moderate correlation with manually scored intra-tumoral stromal TILs in 529 patients for which both were available (Spearman’s ρ = 0.45, *P*<.001; Supplementary Figure 4). When comparing the effect estimates of manually scored TILs with AI-detected TILs for prediction of ORR, AI-detected TILs showed a stronger association per 10% increase (OR 1.48 [95% CI 1.21–1.81] vs. 1.14 [95% CI 1.05–1.23]), indicating a higher predictive value (Table 3).

**Table 3.** Association of AI-detected TILs and manually scored TILs with response, progression free survival and overall survival in 529 patients treated with immune checkpoint inhibition. Results are also shown stratified for 317 patients treated with anti-PD1 monotherapy and 208 patients treated with anti-PD1 + anti-CTLA4. Abbreviations: CI, confidence interval; OR, odds ratio; HR, hazard ratio; ORR, objective response rate; PFS, progression free survival; OS, overall survival

| Outcome | Group | Univariable OR/HR [95% CI] per 10% increase<br>AI-detected TILs | Univariable OR/HR [95% CI] per 10% increase<br>Manual scored TILs |
| --- | --- | --- | --- |
| **ORR** | All | 1.48 [1.21 – 1.81] | 1.14 [1.05 – 1.23] |
| **ORR** | Anti-PD1 | 1.65 [1.28 – 2.13] | 1.18 [1.07 – 1.30] |
| **ORR** | Anti-PD1 + Anti-CTLA4 | 1.23 [0.88 – 1.72] | 1.05 [0.90 – 1.22] |
| **PFS** | All | 0.80 [0.71 – 0.91] | 0.94 [0.90 – 0.99] |
| **PFS** | Anti-PD1 | 0.75 [0.64 – 0.87] | 0.92 [0.87 – 0.98] |
| **PFS** | Anti-PD1 + Anti-CTLA4 | 0.92 [0.75 – 1.14] | 0.99 [0.90 – 1.09] |
| **OS** | All | 0.81 [0.70 – 0.94] | 0.93 [0.87 – 0.98] |
| **OS** | Anti-PD1 | 0.75 [0.63 – 0.90] | 0.91 [0.85 – 0.98] |
| **OS** | Anti-PD1 + Anti-CTLA4 | 0.95 [0.75 – 1.21] | 0.98 [0.88 – 1.10] |

## Discussion

In this multicenter study with a broad and representative cohort, we demonstrate that in patients with metastatic cutaneous melanoma, the presence of AI-detected TILs in pre-treatment metastatic specimens is associated with an improved likelihood of responding to ICI and a lower risk of disease progression and improved overall survival. Importantly, these correlations were independent of known clinical predictors.

The results of our study are in line with the findings of Chatziioannou et al., who reported an association between electronic TILs score and prolonged PFS and melanoma-specific survival in 101 ICI-treated patients with advanced melanoma. In their study, a threshold of 12.2% electronic TILs within manually annotated tumor regions was identified as the optimal prognostic cut-off for PFS. This threshold was significantly associated with prolonged melanoma-specific survival in a Cox regression model adjusted for sex and LDH.^16^ Our study included a substantially larger cohort of 1,202 patients, enabling the assessment of TILs as a continuous variable, which demonstrated an association between higher TIL levels and improved clinical outcomes. In addition, we adjusted for clinical variables known to be associated with response, providing an assessment of the independent predictive value of TILs.

The results of our study and the study performed by Chatziioannou et al. contradict the study performed by Shen et al. who did not find a correlation between TILs and PFS in 144 advanced melanoma patients.^13,14^ However, it is unclear what proportion of samples analyzed in their study were primary tumors versus metastases, as both were included. In our previous work, we found no association between TILs in primary melanomas and response to ICIs, which may partly explain the absence of a correlation. When considering the work of Chatziioannou et al. and our previous work in manual TILs, both of which focused on metastatic samples, there is increasing evidence to support a relationship between TILs in pre-treatment metastasis and treatment outcomes. This is further supported by multiple studies showing associations between intratumoral CD8^+^ and CD4^+^ lymphocyte presence and response to ICIs.^9,28,29^

A major advantage of AI-based TIL quantification is its consistency and reproducibility, whereas interobserver agreement for manual scoring among pathologists is often only moderate.^7,8^ Furthermore, TILs can be quantified on routinely available H&E-stained slides, eliminating the need for additional immune histochemical staining and making it a practical and accessible biomarker in clinical practice. In our study, AI-detected TILs demonstrated stronger associations with clinical outcomes compared to manually scored TILs, further highlighting their potential. To support transparency and reproducibility, our AI model, as well as a part of the developmental dataset, have been made publicly available, enabling further external validation and future clinical implementation.^17,30^

Our study has several strengths, including its size, multicenter design, and the use of histopathology images from 30 institutions with varying staining protocols, which enhances the generalizability and applicability of our findings. Furthermore, we evaluated TILs as a continuous variable rather than relying on an optimal threshold, allowing for a more nuanced analysis of their predictive value.

This study is also subject to several limitations. Some patients were excluded from the analysis due to the absence of available histopathology samples, however no significant differences were observed between included and excluded patients. AI-based segmentation may also occasionally misclassify TILs; however, no major misclassifications of tumor cells or TILs were identified upon visual inspection. In addition, we assessed only a single slide of a metastatic lesion per patient, despite known intra- and interlesional heterogeneity, which may limit the generalizability of findings across all tumor sites.^31^ However, in the single selected sample before therapy initiation there was already predictive value supporting the predictive capability of TILs in advanced melanoma. Lastly, the observed ORR in our cohort was relatively high. This may, in part, be explained by the use of investigator-assessed treatment response in the Dutch Melanoma Treatment Registry. The response rate is similar to earlier studies published from this registry data.^2^

Whereas this study shows that TILs can be used to identify patients with a favorable prognosis, reaching a response rate of 74.6% in patients with high TIL scores (above 34.5%), a major limitation remains that it is not sufficiently possible to identify non-responders as demonstrated by the ORR of 40.7% in patients with less than 3.54% TILs. Future research should therefore focus on improving the negative predictive value of AI-assisted histopathology analysis. Previous studies suggest that not only the quantity but also the spatial context of TILs, such as their presence within the tumor center or at the invasive margin and distance between TILs and tumor cells, may be prognostically relevant ^28,32^. Investigating whether foundation models, capable of directly analyzing whole-slide histopathological images, can outperform current AI-based methods is another promising direction ^33,34^. These models may better capture tumor–immune interactions which are associated with clinical outcomes. However, a downside is their limited interpretability when compared to TIL analysis, as they are based on abstract feature representations of the histopathology image data.

Regarding the current TIL detection model, future efforts should focus on integrating AI-detected TILs into prediction models that can be used in clinical practice, not only to guide the decision to start treatment, but also to de-escalate treatment. TILs appear to be particularly valuable in identifying patients with a high likelihood of response. In this light, the algorithm might be valuable in the neoadjuvant setting, identifying for whom anti-PD1 monotherapy suffices.

## Conclusion

In this large multicenter study, AI-detected TILs in pre-treatment metastatic melanoma specimens are predictive of response to ICI and associated with improved PFS and OS. TILs provide a standardized, reproducible, and potentially more predictive alternative to manual scoring and can be derived from routine H&E-stained slides, making them a practical and accessible biomarker. These findings support the potential of AI-driven histopathological analysis as a predictive tool in melanoma. Further research is needed to improve the predictive value of AI-detected TILs and its use in treatment guidance and de-escalation.

## Data Availability

Part of the developmental dataset used for model training is publicly available on Zenodo 1. The model architecture is available on Github2 and model specific weights are automatically downloaded upon usage. The whole slide images and associated patient outcome data used in this study cannot be shared due to confidentiality agreements and data sharing restrictions related to patient privacy.
1. Schuiveling M. (2025) Melanoma Histopathology Dataset with Tissue and Nuclei Annotations. Published online March 19, 2025. https://zenodo.org/records/15050523
2. Schuiveling M. (2025) HoVer-NeXt inference for TIL quantification in melanoma: Fork of HoVer-NeXt inference by Baumann et al. with additional pipelines and dataset support for melanoma research [Computer software]. Published online May 27, 2025. Available at: https://github.com/mschuiveling/hover-next-inference-tils-melanoma Forked from: Baumann E, Dislich B, Rumberger JL, Nagtegaal ID, Martínez MR, Zlobec I. HoVer-NeXt inference. [Original software]. Available at: https://github.com/digitalpathologybern/hover_next_inference

https://zenodo.org/records/15050523

https://github.com/mschuiveling/hover-next-inference-tils-melanoma

## Acknowledgements

The authors would like to thank PALGA, the nationwide network and registry of histo- and cytopathology in the Netherlands, for providing histopathological data and for their help in the collection of FFPE tissue.

## Data Sharing Statement

Part of the developmental dataset used for model training is publicly available on Zenodo ^17^. The model architecture is available on Github^18^ and model specific weights are automatically downloaded upon usage. The whole slide images and associated patient outcome data used in this study cannot be shared due to confidentiality agreements and data sharing restrictions related to patient privacy.

## Abbreviations

AI: Artificial Intelligence
AJCC: American Joint Committee on Cancer
CI: Confidence Interval
F_1_: score Harmonic mean of precision and recall in model evaluation
H&E: Hematoxylin and Eosin
ICI: Immune Checkpoint Inhibition
IQR: Interquartile Range
LDH: Lactate Dehydrogenase
MICE: Multiple Imputation by Chained Equations
OR: Odds Ratio
ORR: Objective Response Rate
OS: Overall Survival
PFS: Progression-Free Survival
TILs: Tumor-Infiltrating Lymphocytes
WHO: World Health Organization

