## Supplements for "AI-detected tumor-infiltrating lymphocytes for predicting outcomes in anti-PD1 based treated melanoma"

**Supplementary methodology on developmental cohort creation and model training**

*Development cohort*

The independent development cohort consisted of 155 unique primary and 197 unique metastatic melanoma samples. From these samples, regions of interest (ROIs) were manually selected and extracted at 40× magnification (0.23 µm/px) with a size of 1,024 × 1,024 pixels. Of the 352 cases, 76 were consultation cases from referral hospitals or general practitioners, introducing variation in staining protocols. Manual ROI selection ensured the inclusion of diverse cell types and staining artefacts, enhancing the generalizability of the models trained on this dataset. Metastatic samples originated from various anatomical sites, including skin, lymph nodes, brain, lung, and liver. All primary samples originated from the skin.

Cell segmentations for the dataset were created using Hover-Net, a nuclei segmentation model pretrained on the PanNuke dataset ^1,2^. Manual annotation of the nuclei classes was performed by a trained physician (M.S.), using the following categories: tumor, stroma, vascular endothelium, histiocyte, melanophage, lymphocyte, plasma cell, neutrophil, apoptotic and epithelium. All annotations were reviewed and corrected where needed by a dermatopathologist (W.A.M.B.). Part of the dataset has been made publicly available to further support the development of cell detection models ^3^ . Further information on the dataset creation has been previously published ^4^.

*Model training*

The model used in this study is based on the Hover-NeXt neural network architecture, a state-of-the-art approach for cell detection capable of efficiently processing digitized whole slide images^5^. Hover-NeXt was trained using 258 ROIs from primary and metastatic melanoma samples, consisting of 116,429 cells, of which 64,559 were tumor cells and 26,657 were TILs. A three-class classification strategy was applied: tumor cells, TILs (lymphocytes and plasma cells), and other, based on prior research showing that this results in the highest detection performance for lymphocytes ^4^.

The weights of the final model can be found on Zenodo ^6^ and the code to use the model can be found on Github ^7^.

*Model evaluation*

The performance of the AI TIL detection was assessed using the F_1_ score. This metric is defined as the harmonic mean of precision (the proportion of true positives among all predicted positives) and recall (the proportion of true positives correctly identified out of all actual positives ). The F_1_ score ranges from 0 (worst) to 1 (perfect performance), reflecting a balance between precision and recall. Performance was evaluated on the validation dataset, consisting of 45,406 annotated cells across 94 ROIs, and compared to the interobserver agreement based on 12 samples independently annotated by a second pathologist (G.E.B.) ^4^. Confidence intervals for the F_1_ score were determined using bootstrapping.

Data handling and visualization were done in Python (version 3.10), using geopandas (1.0.1), pandas (2.2.2), numpy (1.24.4) and matplotlib (3.9.2) and by use of Qupath^8^

**Supplementary tables**

**Supplementary table 1.** Patient characteristics of included (having a pre-treatment metastatic melanoma specimen available) versus excluded (not having a pre-treatment metastatic melanoma specimen available) patients. Abbreviations: IQI, interquartile interval; LDH, Lactate dehydrogenase; TIL, tumor-infiltrating lymphocytes; ULN, upper limit of normal; WHO, World Health Organization.

|  |  | Grouped by Therapy | | |
| --- | --- | --- | --- | --- |
|  |  | All patients | Included | Excluded |
| n |  | 1891 | 1202 | 689 |
| Age, median [Q1,Q3] |  | 65.1 (13.0) | 64.9 (12.8) | 65.5 (13.5) |
| Sex, n (%) | Female | 726 (38.4) | 445 (37.0) | 281 (40.8) |
|  | Male | 1165 (61.6) | 757 (63.0) | 408 (59.2) |
| WHO performance status, n (%) | WHO 0 | 1011 (53.5) | 640 (53.2) | 371 (53.8) |
|  | WHO 1 | 652 (34.5) | 409 (34.0) | 243 (35.3) |
|  | WHO 2 or above | 135 (7.1) | 85 (7.1) | 50 (7.3) |
|  | Missing | 93 (4.9) | 68 (5.7) | 25 (3.6) |
| Disease Stage (AJCC 8th Edition), n (%) | IIIC | 172 (9.1) | 109 (9.1) | 63 (9.1) |
|  | M1a | 163 (8.6) | 102 (8.5) | 61 (8.9) |
|  | M1b | 289 (15.3) | 178 (14.8) | 111 (16.1) |
|  | M1c | 822 (43.5) | 529 (44.0) | 293 (42.5) |
|  | M1d with non- symptomatic brain metastasis | 269 (14.2) | 180 (15.0) | 89 (12.9) |
|  | M1d with symptomatic brain metastasis | 176 (9.3) | 104 (8.7) | 72 (10.4) |
| BRAF V600E/K Mutation, n (%) | Wildtype | 1072 (56.7) | 675 (56.2) | 397 (57.6) |
|  | Mutant | 618 (32.7) | 402 (33.4) | 216 (31.3) |
|  | Missing | 201 (10.6) | 125 (10.4) | 76 (11.0) |
| LDH levels, n (%) | Normal | 1296 (68.5) | 832 (69.2) | 464 (67.3) |
|  | 1-2 x ULN | 432 (22.8) | 272 (22.6) | 160 (23.2) |
|  | >2 x ULN | 141 (7.5) | 85 (7.1) | 56 (8.1) |
|  | Missing | 22 (1.2) | 13 (1.1) | 9 (1.3) |
| Therapy | Anti-PD1 | 1216 (64.3) | 779 (64.8) | 437 (63.4) |
|  | Anti-PD1 + anti-CTLA4 | 675 (35.7) | 423 (35.2) | 252 (36.6) |
| Objective response rate, n (%) | Yes | 1036 (54.8) | 667 (55.5) | 369 (53.6) |
|  | No | 813 (43.0) | 510 (42.4) | 303 (44.0) |
|  | Missing | 42 (2.2) | 25 (2.1) | 17 (2.5) |
| Progression free survival, months | Median (95% CI) | 9.0 [95% CI 7.9 - 10.2] | 9.0 [95% CI 7.9 - 10.2] | 9.2 [95% CI 6.9 - 11.8] |
| Overall survival, months | Median (95% CI) | 35.1 [95% CI 31.1 - 41.6] | 35.0 [95% CI 30.5 - 42.3] | 36.7 [95% CI 29.0 - 53.7] |

**Supplementary table 2.** Average tumor-infiltrating lymphocyte (TIL) percentage per organ. Abbreviations: TIL; tumor-infiltrating lymphocyte

| Metastasis Location | TIL Percentage (%), Median [Min, Max] | Number of Samples |
| --- | --- | --- |
| Lymph node | 12.84% [1.22% - 69.41%] | 390 |
| Skin | 8.01% [0.48% - 64.2%] | 342 |
| Soft Tissue | 10.71% [2.09% - 38.74%] | 113 |
| Lung | 11.32% [0.44% - 53.5%] | 84 |
| Liver | 7.27% [0.75% - 26.88%] | 65 |
| Brain | 8.54% [1.48% - 66.29%] | 56 |
| Gastro-intestinal organ | 7.26% [1.76% - 35.02%] | 46 |
| Bone | 7.26% [1.52% - 52.31%] | 26 |
| Other | 7.08% [4.43% - 40.95%] | 8 |
| Adrenal gland | 6.52% [1.14% - 21.09%] | 7 |
| Urogenital organ | 13.09% [7.65% - 18.58%] | 6 |
| Gall bladder | 12.51% [7.1% - 15.13%] | 3 |
| Unknown | 6.29% [0.3% - 32.31%] | 56 |

**Supplementary Table 3.** Full multivariable logistic and Cox regression analyses in all patients. Results shown as odds ratios and hazard ratios with 95% confidence intervals. Abbreviations CI, confidence interval; OR, odds ratio; HR, hazard ratio; ORR, objective response rate; PFS, progression free survival; OS, overall survival.

| Variable | OR [95% CI] for ORR | HR [95% CI] for PFS | HR [95% CI] for OS |
| --- | --- | --- | --- |
| TILs (per 10% increase) | 1.40 [1.23 - 1.59] | 0.85 [0.79 - 0.92] | 0.83 [0.76 - 0.91] |
| Age (per year) | 1.01 [1.00 - 1.02] | 1.00 [0.99 - 1.00] | 1.01 [1.00 - 1.02] |
| Male sex | 0.94 [0.73 - 1.22] | 1.06 [0.91 - 1.23] | 1.03 [0.86 - 1.23] |
| WHO PS 1 vs 0 | 0.68 [0.51 - 0.89] | 1.07 [0.91 - 1.25] | 1.09 [0.90 - 1.31] |
| WHO PS ≥2 vs 0 | 0.42 [0.25 - 0.70] | 1.68 [1.28 - 2.20] | 2.25 [1.67 - 3.04] |
| LDH 1-2× ULN vs normal | 0.82 [0.60 - 1.12] | 1.22 [1.02 - 1.45] | 1.27 [1.04 - 1.56] |
| LDH >2 ULN vs normal | 0.31 [0.18 - 0.54] | 2.17 [1.65 - 2.86] | 2.86 [2.12 - 3.87] |
| BRAF V600E mutant vs wildtype | 0.84 [0.63 - 1.11] | 1.12 [0.95 - 1.31] | 0.83 [0.69 - 1.01] |
| Anti-PD1 + Anti-CTLA4 vs Anti-PD1 | 1.87 [1.39 - 2.52] | 0.70 [0.59 - 0.83] | 0.73 [0.60 - 0.90] |
| Stage M1a vs IIIC | 1.76 [0.97 - 3.19] | 0.95 [0.67 - 1.35] | 0.95 [0.59 - 1.53] |
| Stage M1b vs IIIC | 1.93 [1.15 - 3.27] | 0.94 [0.69 - 1.29] | 1.20 [0.80 - 1.80] |
| Stage M1c vs IIIC | 1.00 [0.64 - 1.56] | 1.19 [0.90 - 1.58] | 1.62 [1.12 - 2.32] |
| Stage M1d (non-symptomatic) vs IIIC | 0.98 [0.58 - 1.65] | 1.44 [1.05 - 1.97] | 2.13 [1.43 - 3.17] |
| Stage M1d (symptomatic) vs IIIC | 0.56 [0.31 - 1.01] | 1.86 [1.31 - 2.62] | 2.86 [1.86 - 4.37] |

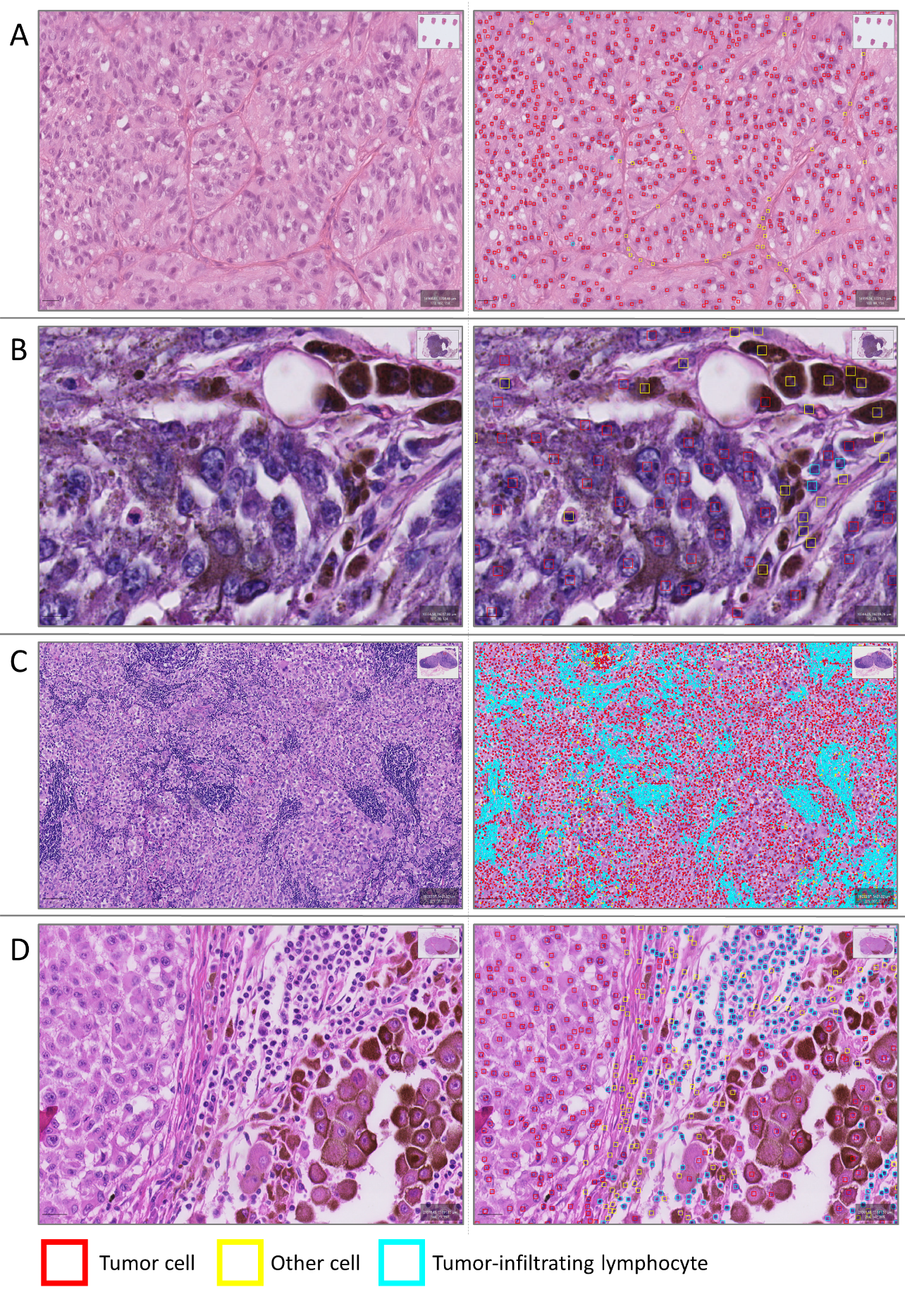
**Supplementary figures**

**Supplementary Figure 1.** Shown in the panels are a digitized zoomed-in region of a melanoma biopsy from a skin metastasis with low presence of tumor-infiltrating lymphocytes (TILs) (A), an excised lymph node metastasis with tumor cells and melanophages labeled as “other” (B), a zoomed-out view of a lymph node metastasis with a high average TIL percentage of 61% (C), and a zoomed-in region of interest with intralesional heterogeneity and variation in TIL density (D).

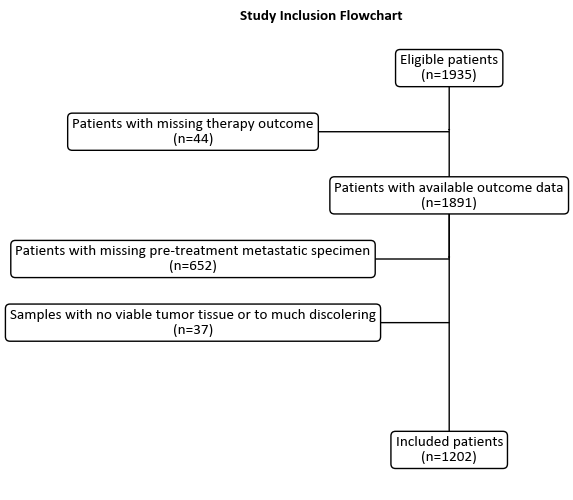

**Supplementary Figure 2.** Flowchart of study population

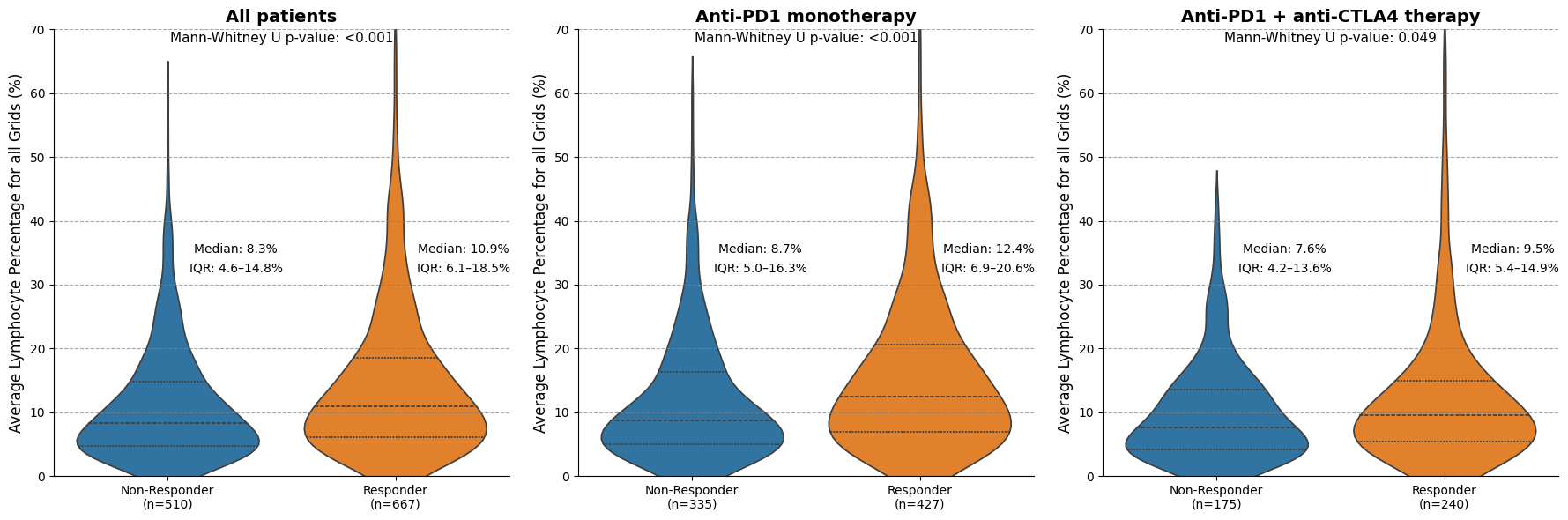
**Supplementary Figure 3.** Comparison of AI-detected TILs in patients stratified by response to immune checkpoint inhibition. The figure includes all patients (n = 1177), those treated with anti-PD1 monotherapy (n = 762), and those treated with anti-PD1 + anti-CTLA4 combination therapy (n = 415). Response assessment data were unavailable for 25 patients. Differences in TIL levels between responders and non-responders were assessed using the Mann–Whitney U test. Across all groups, responders showed higher TIL levels compared to non-responders.

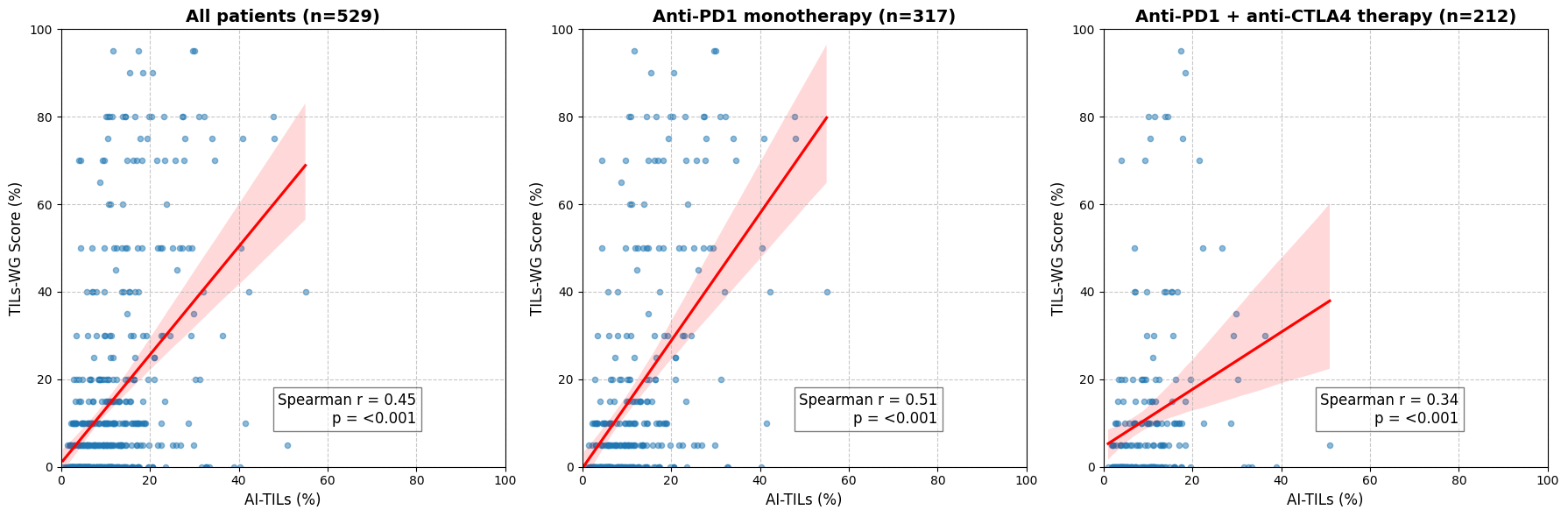

**Supplementary Figure 4.** Correlation between AI-detected tumor-infiltrating lymphocytes and manually scored stromal TILs (scored according to the guidelines of the TIL working group) in all patients, monotherapy patients, and patients treated with combination therapy (anti-PD1+ anti-CTLA4). Spearman correlation coefficients and p-values are displayed in each panel, showing a moderate correlation in all patients, which is stronger in monotherapy-treated patients.
